# Determination of IgG1 and IgG3 SARS-CoV-2 spike protein and nucleocapsid binding – Who is binding who and why?

**DOI:** 10.1101/2021.06.17.21259077

**Authors:** Jason K IIes, Raminta Zmuidinaite, Christoph Sadée, Anna Gardiner, Jonathan Lacey, Stephen Harding, Gregg Wallis, Roshani Patel, Debra Roblett, Jonathan Heeney, Helen Baxendale, Ray K Iles

## Abstract

The involvement of IgG3 in the humoral immune response to SARS-CoV2 infection has been implicated in the pathogenesis of ARDS in COVID-19. The exact molecular mechanism is unknown but may be due to the differential ability of IgG3 Fc region to fix complement and stimulate cytokine release. We examined convalescent patients’ antibodies binding to immobilised nucleocapsid and spike protein by MALDI-ToF mass spectrometry. IgG3 was a major immunoglobulin found in all samples. Differential analysis of the spectral signatures found for nucleocapsid versus spike protein demonstrated that the predominant humoral immune response to nucleocapsid was IgG3, whilst against spike it was IgG1. However, the spike protein displayed a strong affinity for IgG3 itself which it would bind from control plasma samples as well as from those previously infected with SARS-CoV2, much in the way Protein-G binds IgG1. Furthermore, detailed spectral analysis indicated a mass shift consistent with hyper-glycosylation or glycation was a characteristic of the IgG3 captured by the spike protein.

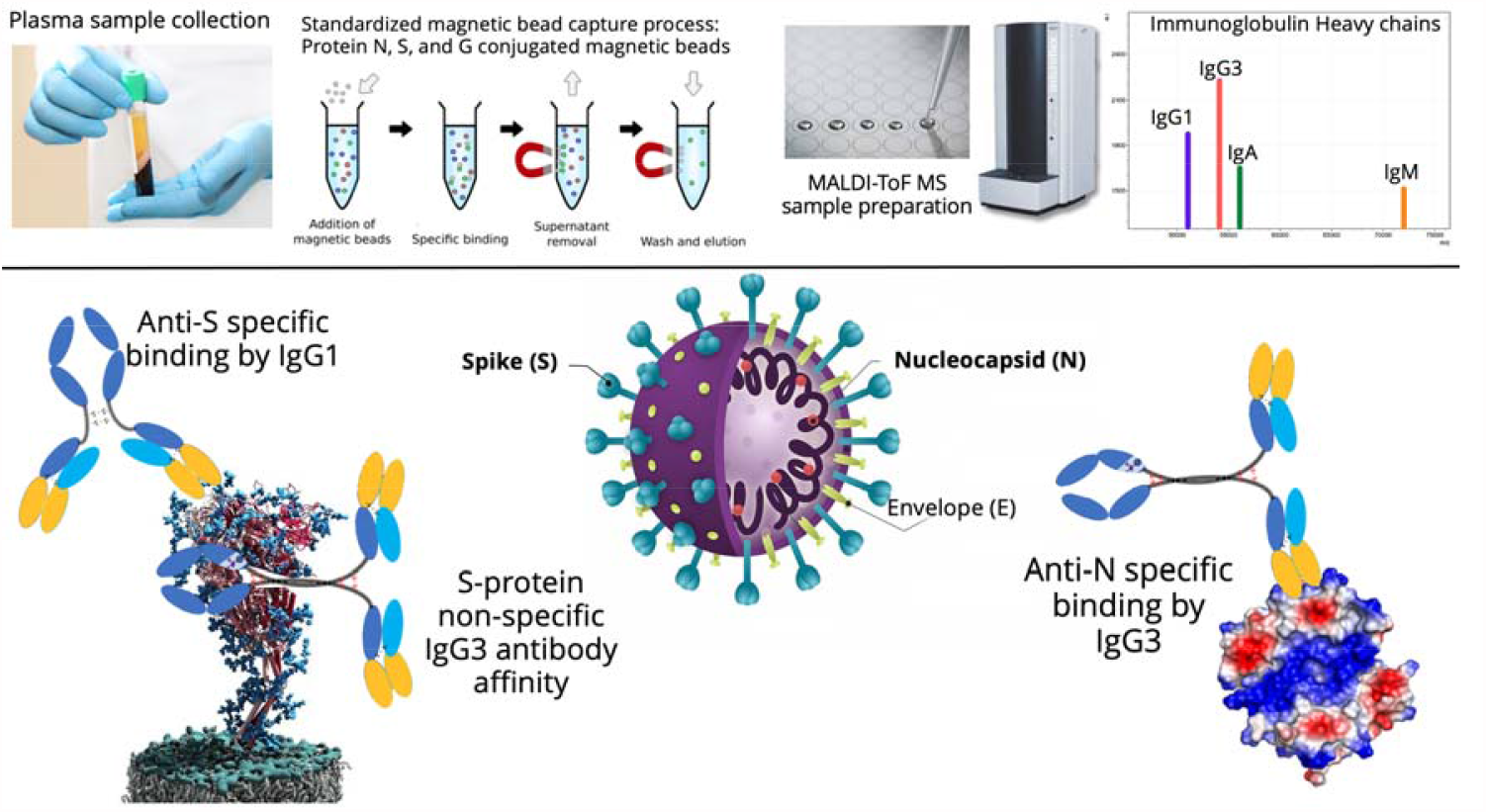

## Introduction

The rapidly developing COVID-19 disease syndrome is a delayed event following initial SARS-CoV-2 infection of the upper respiratory tract followed by lower respiratory tissue involvement and a progressive hyper-inflammatory immune response [1]. Somewhat counter intuitively the marked feature of those requiring hospital treatment is the extremely high titres of anti-viral antibodies and cytokine elevation in those with severe COVID-19 disease at time of hospitalisation, associated with high viral load [2, 3]. However, despite this immune response, there is an invasion of alveolar cells and the blood stream by viral particles [4].

The humoral response to SARS-CoV-2 is primarily directed towards the nucleocapsid (N-protein), and the spike protein (S-protein) complex [5]. By the stage of the onset of acute respiratory distress syndrome (ARDS), the initial IgM antibody response to the virus has declined and is replaced by IgA and IgG antibodies. IgG is the most abundant class of antibody found in the convalescent plasma of those recovering from COVID-19 ARDS [6]; and the onset of ARDS appears to correspond with the time of antibody class switch to IgG [7] (see Figure 1 panel A). Antibody responses directed at the spike protein and the receptor binding domain (RBD) in particular, have been identified as the main neutralizing component of the SARS-CoV-2 antibody response [8,9,10]. A distinct antibody signature has been linked to different COVID-19 disease outcomes: early spike-specific responses were associated with a positive outcome, while early nucleocapsid specific responses were associated with a negative outcome and death. The differential impact of Fc-associated functions of the antibody response such as antibody-mediated phagocytosis, cytotoxicity, and complement deposition which are critical for disease resolution are less well understood[11].

**Figure 1.**
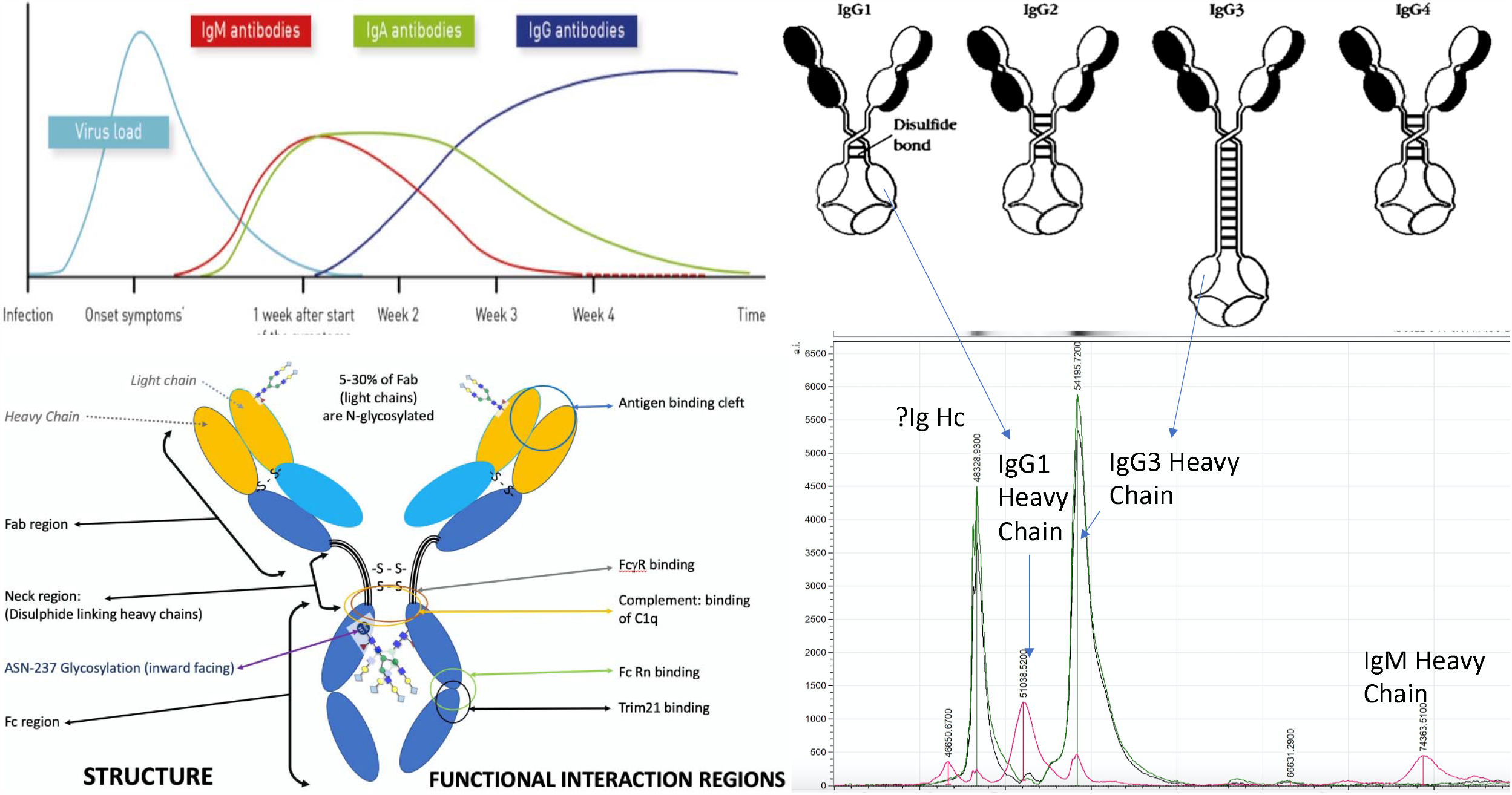
A schematic of the Humoral response - immunoglobulin major class switching following SARS-CoV2 infection and COVID-19 progresses, is illustrated in panel A. IgG predominates in 3-4 weeks after onset of symptoms and the major structural and functional domains of IgG are also illustrated. The variation in the neck region of the 4 subtypes of human IgG are shown in panel B. IgG3 is the largest and its heavy chain resolution in MALDI-ToF mass spectra is indicated at 54,000m/z (IgG3 Hc). Also indicated is IgG1 heavy chain (IgG1 Hc) which resolves at 51,000m/z. The position of IgM heavy chain peak (IgM Hc) is indicated at 74,000m/z. An as yet to be fully identified Ig (IgX Hc), thought to be either IgG2 or IgG4, found in patient samples is shown at 49,000m/z.

IgG consists of four sub classes each defined by structural differences within and adjacent to the hinge region associated with the constant region of the antibody (Fc). This effect Fc receptor binding and complement activation function (see Figure 1 panel b and Table 1). A highly elevated disproportionate IgG subclass response, dominated by IgG3, has been implicated as a discriminant marker of adverse outcome in COVID-19 patients [12].

**Table 1.**
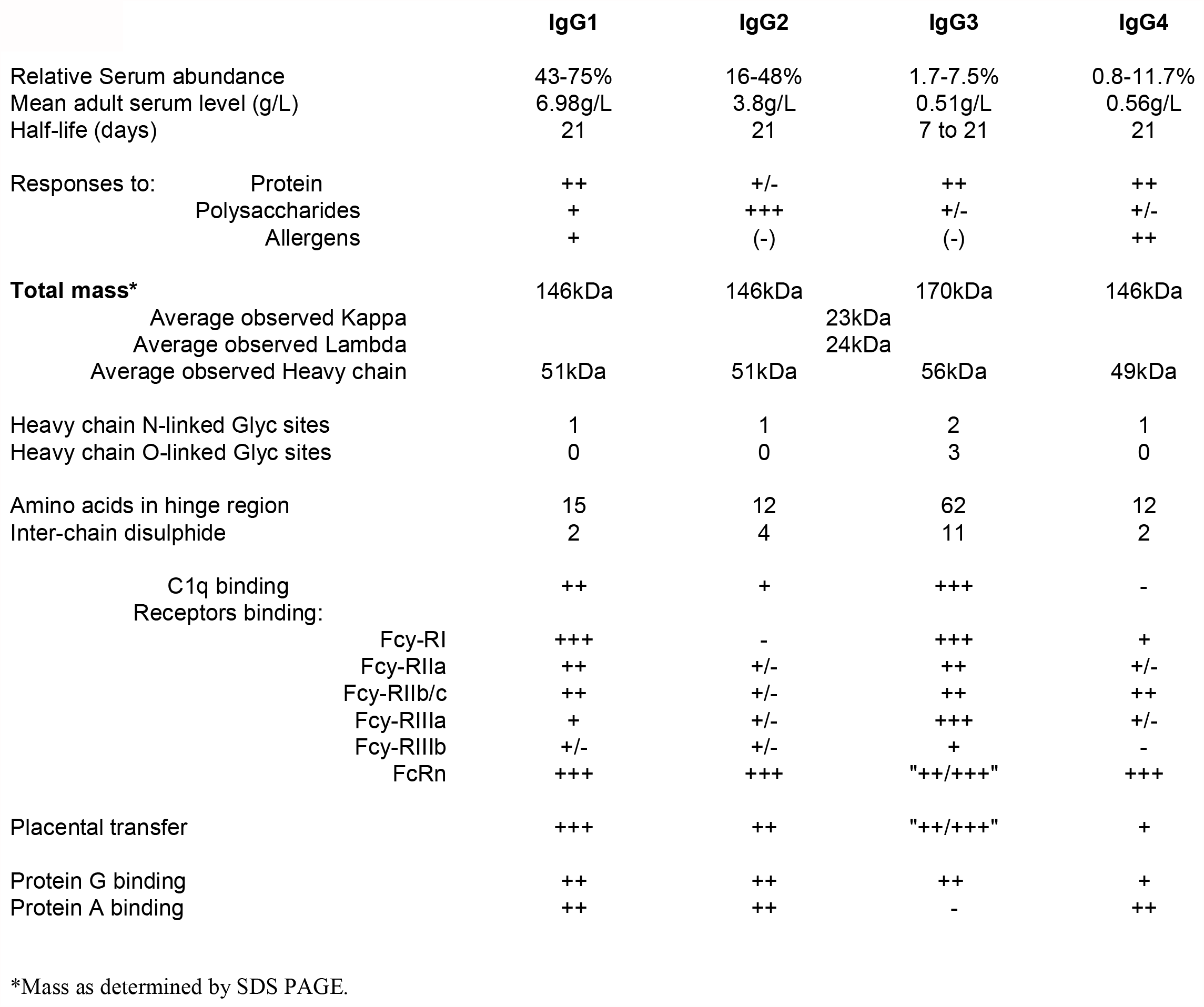
Descriptive comparison of physical-structure and functional-biology differences between the four human IgG subtypes.

In this study, binding proteins, in particular antibodies, were captured from COVID-19 convalescent patients’ plasma samples and examined by MALDI-ToF mass spectrometry. The mass spectral signatures of Immunoglobulin (Ig) class and IgG subclass were determined, in particular, that of the respective heavy chains of Immunoglobulins, and quantified (Figure 1 panel D). Here we report the comparison of IgG1 and IgG3 captured by immobilised N and S-proteins in relation to disease severity post SARS-CoV2 infection.

## Materials and methods

### Samples

Serum and plasma samples were obtained from Health care workers (HCWs) and patients referred to the Royal Papworth Hospital for critical care. COVID-19 patients hospitalised during the first wave and as well as NHS healthcare workers working at the Royal Papworth Hospital in Cambridge, UK served as the exposed HCW cohort (Study approved by Research Ethics Committee Wales, IRAS: 96194 12/WA/0148. Amendment 5). NHS HCW participants from the Royal Papworth Hospital were recruited through staff email over the course of 2 months (20^th^ April 2020-10^th^ June 2020) as part of a prospective study to establish seroprevalence and immune correlates of protective immunity to SARS-CoV-2. Patients were recruited in convalescence either pre-discharge or at the first post-discharge clinical review. All participants provided written, informed consent prior to enrolment in the study. Sera from NHS HCW and patients were collected between July and September 2020, approximately 3 months after they were enrolled in the study.

For cross-sectional comparison, representative convalescent serum and plasma samples from sero-negative HCWs, sero-positive HCW and convalescent PCR-positive COVID-19 patients were obtained. The serological screening to classify convalescent HCW as positive or negative was done according to the results provided by a CE-validated Luminex assay detecting N-, RBD- and S-specific IgG, a lateral flow diagnostic test (IgG/IgM) and an Electro-chemiluminescence assay (ECLIA) detecting N- and S-specific IgG. Any sample that produced a positive result by any of these assays was classified as positive. Thus, the panel of convalescent plasma samples (3 months post-infection) were grouped in three categories: and A) Sero-negative Staff (n=30 samples). B) Sero-positive Staff (n=31 samples); C) Patients (n=38 samples) [13].

### Antigen coupled magnetic beads

Protein-G coupled magnetic beads were purchased from Cytivia Ltd (Amersham Place, Little Chalfont, Buckinghamshire, UK). Recombinant nucleocapsid and recombinant stabilized complete spike protein magnetic beads were made by Bindingsite Ltd (Birmingham, UK).

The viral spike protein (S-protein) is present on virions as pre-fusion trimers with the receptor binding domain of the S1 region stochastically open or closed, an intermediary where the S1 region is cleaved and discarded, and the S2 undergoes major confirmation changes to expose and then retract its fusion peptide domain [14]. Here the S-protein was modified to disable the S1/S2 cleavage site and maintain the pre-fusion stochastic confirmation [15].

### Semi-automated magnetic bead capture processing

The processes of magnetic bead capture, washing, agitation and target binding protein elution can vary dramatically due to damage from too vigorous mixing and yet insufficient washing can result in large amounts of non-specific binding proteins being recovered. To minimise these problems, and individual operator variability, in the efficiency of target binding proteins recoveries, the Crick automated magnetic rack system was employed. This has been described in full previously [16].

### Pre-processing of the magnetic beads

1.5µl microcentrifuge tubes were loaded into the automated magnetic rack. Protein G (GE), purified nucleocapsid or purified stabilized complete spike magnetic beads (Bindingsite, Birmingham UK) in their buffer solutions were vortexed to ensure an even distribution of beads within the solution. 10µl of the appropriate magnetic beads were pipetted into each tube. 100µl of wash buffer, 0.1% Tween 20 in Dulbecco’s phosphate, buffered saline (DPBS) was pipetted into each tube before resuspending. After mixing for several minutes, the instrument pulled the antigen beads to one side allowing the wash buffer to be carefully discarded. The wash cycle was repeated three times.

### Sample processing and binding fraction elution from the magnetic beads

45µl of 10X DPBS was pipetted into each of the tubes containing the washed magnetic beads. 5µl of vortexed neat plasma was pipetted and pump mixed into a tube containing the beads, repeating for each plasma sample. After the resuspension, and automated mixing for 20 minutes, the beads were magnetically collected to one side and the non-bound sample discarded. A further 3 wash cycles were conducted using 0.1% DPBS. Subsequently, another 3 wash cycles were conducted after this, using ultra-pure water, discarding the water after the last cycle. 15µl of recovery solution (20mM tris(2-carboxyethyl)phosphine (TCEP) (Sigma-Aldrich, UK) + 5% acetic acid + ultra-pure water) was pipetted into the tubes. The tubes were run alternatively between the ‘Resuspend’ and ‘Mix’ setting for several minutes. After pulling the extracted magnetic beads to one side the recovery solution was carefully removed using a pipette and placed into a clean, labelled 0.6µl microcentrifuge tube. This recovery solution was the eluant from the beads and contained the desired proteins.

### Sample Analysis by MALDI-ToF Mass spectrometry

Mass spectra were generated using a 15mg/ml concentration of sinapinic acid (SA) matrix. The elute from the beads was used to plate with no further processing. 1µl of the eluted samples were taken and plated on a 96 well stainless-steel target plate using a sandwich technique. The MALDI-ToF mass spectrometer (microflex® LT/SH, Bruker, Coventry, UK) was calibrated using a 2-point calibration of 2mg/ml bovine serum albumin (33,200 m/z and 66,400 m/z) (Pierce™, ThermoFisher Scientific). Mass spectral data were generated in a positive linear mode. The laser power was set at 65% and the spectra were generated at a mass range between 10,000 to 200,000 m/z; pulsed extraction set to 1400ns.

A square raster pattern consisting of 15 shots and 500 positions per sample was used to give 7500 total profiles per sample. An average of these profiles was generated for each sample, giving a reliable and accurate representation of the sample across the well. The raw, averaged spectral data was then exported in a text file format to undergo further mathematical analysis.

### Spectral Data processing

Mass spectral data generated by the MALDI-ToF instrument were uploaded to an open-source mass spectrometry analysis software mMass(tm) [17], where it was processed by using; a single cycle, Gaussian smoothing method with a window size of 300 m/z, and baseline correction with applicable precision and relative offset depending on the baseline of each individual spectra. In software, an automated peak-picking was applied to produce a peak list which was then tabulated and used in subsequent statistical analysis.

### Statistical analysis

Peak mass and peak intensities were tabulated in excel and plotted in graphic comparisons of distributions for each antigen capture and patient sample group. Means and Medians were calculated and, given the asymmetric distributions found, non-parametric statistics were applied, such as Mann Whitney U test, when comparing differences in group distributions.

## Results

Post elution from the respective antigen coupled magnetic beads MALDI-ToF mass spectra was obtained and peaks recorded. These were matched against reference MALDI-ToF mass spectra of preparation of purified human serum proteins run under the same reducing and acetic acid pH conditions: human serum albumin, Transferrin (Merck Life Science UK Ltd, Dorset, England), IgG1, IgG3, IgA & IgM (Abcam, Discovery Drive, Cambridge, Biomedical Campus, Cambridge, UK).

From pooled polyclonal immunoglobulin isolate, IgG1 heavy chains averaged at peak apex ∼51,000m/z, IgG3 at ∼54,000m/z, IgA at ∼56,000m/z and IgM at ∼74,000m/z. Human Albumin was at 66,400m/z (1+) and Transferrin at ∼79,600m/z. All are broad heterogenous peaks reflecting glycosylation/glycation and sequence variation. In the protein-G, N- and S-protein magnetic bead isolates, the dominance of IgG1 was almost always lost giving way to IgG3 peak prominence at ∼54,000m/z and a peak at ∼49,000m/z. The latter has revealed IgG common peptide sequence fragment matches, following formic acid lysis, but a precise identity as IgG4 or IgG2 has yet to be confirmed (unpublished data) (see Figure 1 panel D). IgA peaks at ∼56,000m/z and IgM at ∼74,000m/z could be detected in some samples but was always minor (at least 1/20^th^) to the IgG subtypes peak intensities. This study focuses on the IgG1 and IgG3 binding to the antigens.

### IgG1 levels and molecular mass

Looking at overall binding Protein G magnetic bead bound large amounts of IgG1 from all samples (peak detected in 96% of samples, median intensity 2870AU), N-protein very little (peak detected in 33% of samples, median intensity 0AU) and S-protein a smaller but significant quantity (peak detected in 72% of samples, median intensity 283AU) (see Figure 2). Differentiating the three sample groups, Protein-G had bound equally high amounts of IgG1 from sero-negative HCW, sero-positive HCW with mild symptoms and COVID-19 ARDS patient samples (see Figure 3). Thus, reflecting the non-specific nature of binding of IgG1 by protein-G. N-protein magnetic beads displayed equally low IgG1 levels in all three sample groups. Thus, there were no significant anti nucleocapsid IgG1 antibodies, nor specific binding of IgG1 by N-protein. Prefusion complete S-protein binding profiles were distinct in the three cohort. S-protein displayed the lowest binding of IgG1 in the sero-negative HCW samples (peak detected in 60% of samples, median intensity 186AU), higher levels in the sero-positive HCW (peak detected in 61% of samples, median intensity 283AU) and significantly higher levels in the COVID-19 ARDS patient samples (peak detected in 92% of samples, median intensity 507AU) (Figure 3 & Table 2 panel A). Thus, reflecting specific anti-spike IgG1 binding to the immobilised spike protein.

**Figure 2.**
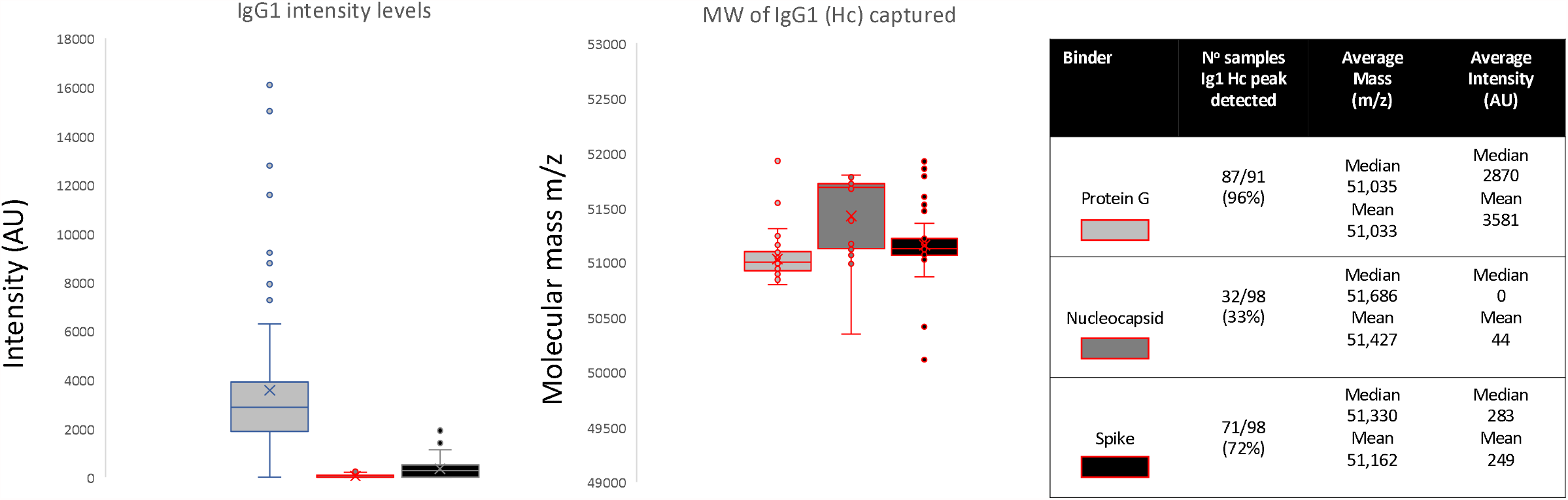
Relative intensities and variance in peak apex molecular mass of IgG1 heavy chains (IgG1 Hc) recovered from the same samples by Protein G, nucleocapsid and stabilised spike protein.

**Figure 3.**
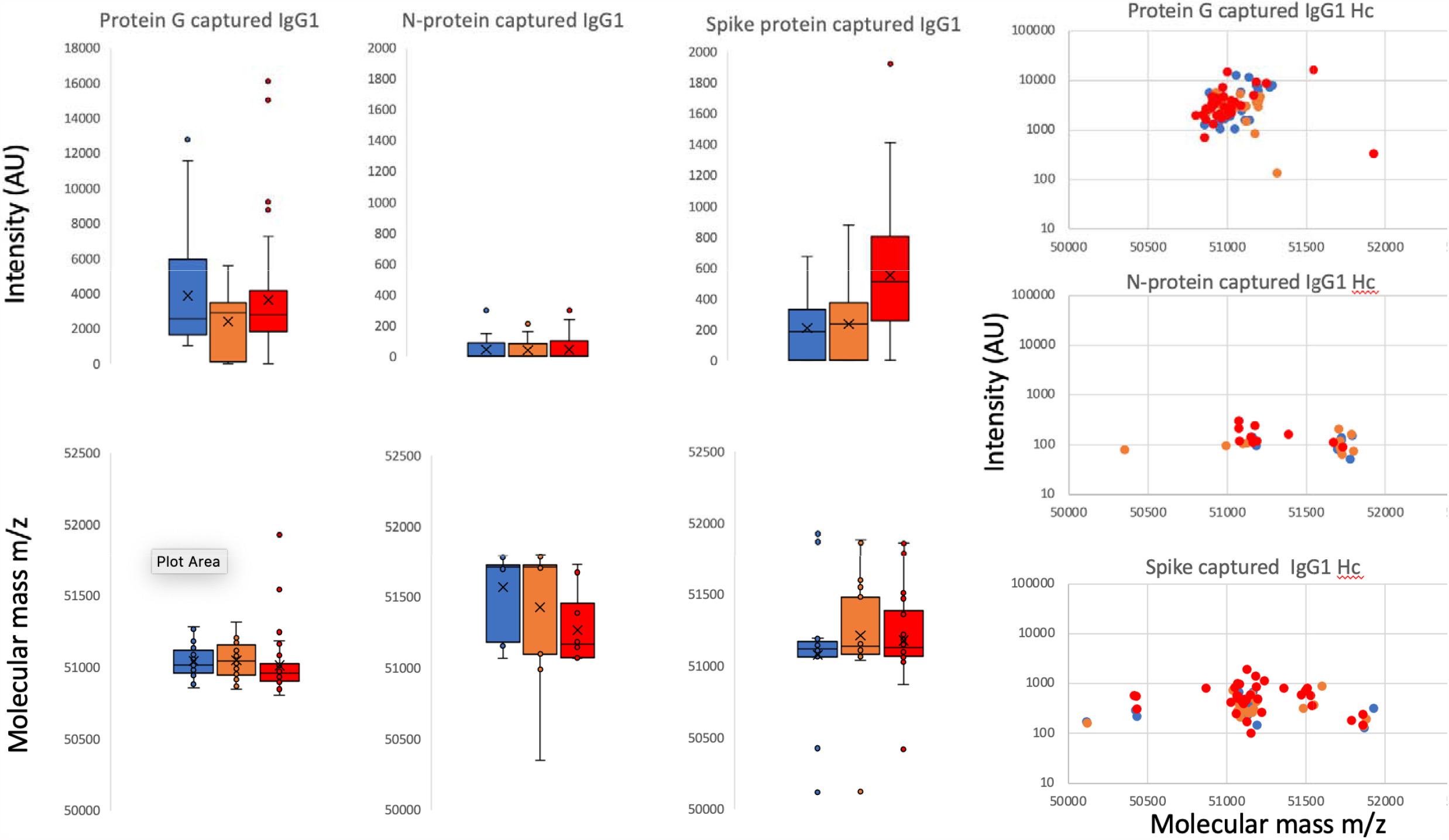
Distribution of intensity for captured and eluted IgG 1 heavy chains (IgG1 Hc) and the relative peak molecular mass being bound. The dot plots to the right are intensity versus molecular mass for individual samples; Blue represents data from SARS-CoV2 sero-negative HCW, Orange from SARS-CoV2 HCW sero-positive having recovered from with mild symptoms and Red sample data from convalescent patients recovering from COVID-19 ARDS.

**Table 2.**
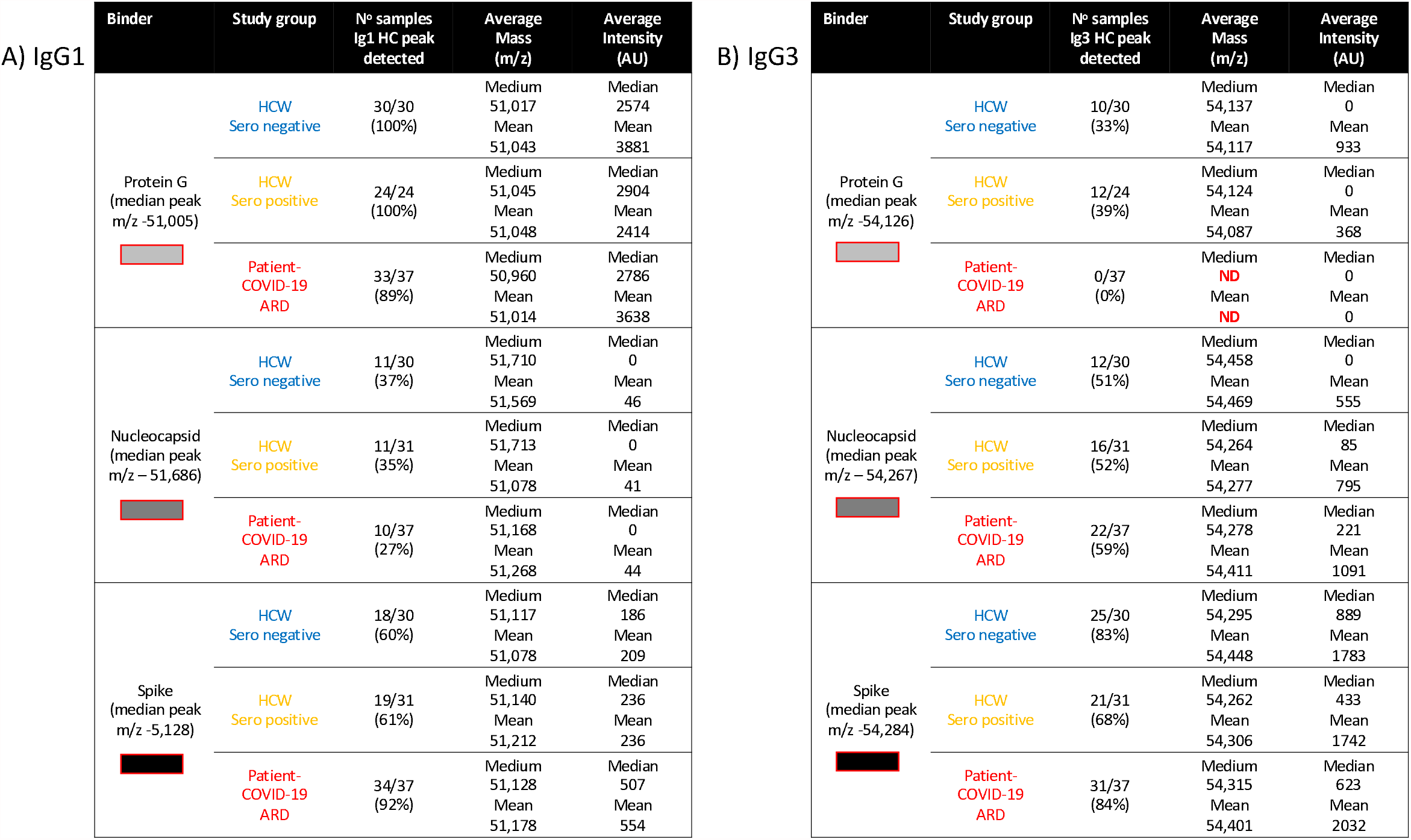
Comparative table of IgG1 and IgG3 spectral analysis: Detection frequency, intensity and molecular mass, for all samples captured by Protein-G 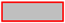, nucleocapsid 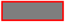 and prefusion complete spike protein 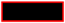. Further delineation is by sample infection status: Blue represents data from SARS-CoV2 sero-negative HCW, Orange from SARS-CoV2 HCW sero-positive having recovered from with mild symptoms and Red sample data from convalescent patients recovering from COVID-19 ARDS.

A) IgG1
| Binder | Study group | N° samples<br>Ig1 HC peak<br>detected | Average<br>Mass<br>(m/z) | Average<br>Intensity<br>(AU) |
| --- | --- | --- | --- | --- |
| Protein G<br>(median peak<br>m/z -51,005) | HCW<br>Sero negative | 30/30<br>(100%) | Medium<br>51,017<br>Mean<br>51,043 | Median<br>2574<br>Mean<br>3881 |
|  | HCW<br>Sero positive | 24/24<br>(100%) | Medium<br>51,045<br>Mean<br>51,048 | Median<br>2904<br>Mean<br>2414 |
|  | Patient-COVID-19<br>ARD | 33/37<br>(89%) | Medium<br>50,960<br>Mean<br>51,014 | Median<br>2786<br>Mean<br>3638 |
| Nucleocapsid<br>(median peak<br>m/z -51,686) | HCW<br>Sero negative | 11/30<br>(37%) | Medium<br>51,710<br>Mean<br>51,569 | Median<br>0<br>Mean<br>46 |
|  | HCW<br>Sero positive | 11/31<br>(35%) | Medium<br>51,713<br>Mean<br>51,078 | Median<br>0<br>Mean<br>41 |
|  | Patient-COVID-19<br>ARD | 10/37<br>(27%) | Medium<br>51,168<br>Mean<br>51,268 | Median<br>0<br>Mean<br>44 |
| Spike<br>(median peak<br>m/z -5,128) | HCW<br>Sero negative | 18/30<br>(60%) | Medium<br>51,117<br>Mean<br>51,078 | Median<br>186<br>Mean<br>209 |
|  | HCW<br>Sero positive | 19/31<br>(61%) | Medium<br>51,140<br>Mean<br>51,212 | Median<br>236<br>Mean<br>236 |
|  | Patient-COVID-19<br>ARD | 34/37<br>(92%) | Medium<br>51,128<br>Mean<br>51,178 | Median<br>507<br>Mean<br>554 |

B) IgG3
| Binder | Study group | N° samples<br>Ig3 HC peak<br>detected | Average<br>Mass<br>(m/z) | Average<br>Intensity<br>(AU) |
| --- | --- | --- | --- | --- |
| Protein G<br>(median peak<br>m/z -54,126) | HCW<br>Sero negative | 10/30<br>(33%) | Medium<br>54,137<br>Mean<br>54,117 | Median<br>0<br>Mean<br>933 |
|  | HCW<br>Sero positive | 12/24<br>(39%) | Medium<br>54,124<br>Mean<br>54,087 | Median<br>0<br>Mean<br>368 |
|  | Patient-COVID-19<br>ARD | 0/37<br>(0%) | Medium<br>ND<br>Mean<br>ND | Median<br>0<br>Mean<br>0 |
| Nucleocapsid<br>(median peak<br>m/z -54,267) | HCW<br>Sero negative | 12/30<br>(51%) | Medium<br>54,458<br>Mean<br>54,469 | Median<br>0<br>Mean<br>555 |
|  | HCW<br>Sero positive | 16/31<br>(52%) | Medium<br>54,264<br>Mean<br>54,277 | Median<br>85<br>Mean<br>795 |
|  | Patient-COVID-19<br>ARD | 22/37<br>(59%) | Medium<br>54,278<br>Mean<br>54,411 | Median<br>221<br>Mean<br>1091 |
| Spike<br>(median peak<br>m/z -54,284) | HCW<br>Sero negative | 25/30<br>(83%) | Medium<br>54,295<br>Mean<br>54,448 | Median<br>889<br>Mean<br>1783 |
|  | HCW<br>Sero positive | 21/31<br>(68%) | Medium<br>54,262<br>Mean<br>54,306 | Median<br>433<br>Mean<br>1742 |
|  | Patient-COVID-19<br>ARD | 31/37<br>(84%) | Medium<br>54,315<br>Mean<br>54,401 | Median<br>623<br>Mean<br>2032 |

Examining the average molecular mass of the IgG1 bound by or binding to the magnetic beads, although a wide variation of mass (+/- 800m/z) could be found between individual sample (Figure 2, Table 2 panel A); no significant patterns could be detected as characteristic of any of the sample groups.

### IgG3 levels and molecular mass

Overall binding showed that Protein-G could bind IgG3 very rarely (peak detected in 24% of samples, median intensity 0AU), compared to that of N-protein (peak detected in 51% of samples, median intensity 103AU) and prefusion S-protein (peak detected in 79% of samples, median intensity 630AU).

Although IgG3 is reported to be bound by Protein-G, in our samples this was low. Furthermore, Protein-G almost completely failed to bind IgG3 in COVID-19 ARDS convalescent patient plasma (peak detected in 0 samples) and only in sero-negative (peak detected in 33% of samples) and sero-positive HCW (peak detected in 39% of samples). Furthermore, the molecular mass of the IgG3 captured by Protein-G was uniform and consistent at 54,124 – 54,137m/z and never captured the larger IgG3 seen in N and S protein eluants (see Figures 4&5 and Table 2 panel B).

**Figure 4.**
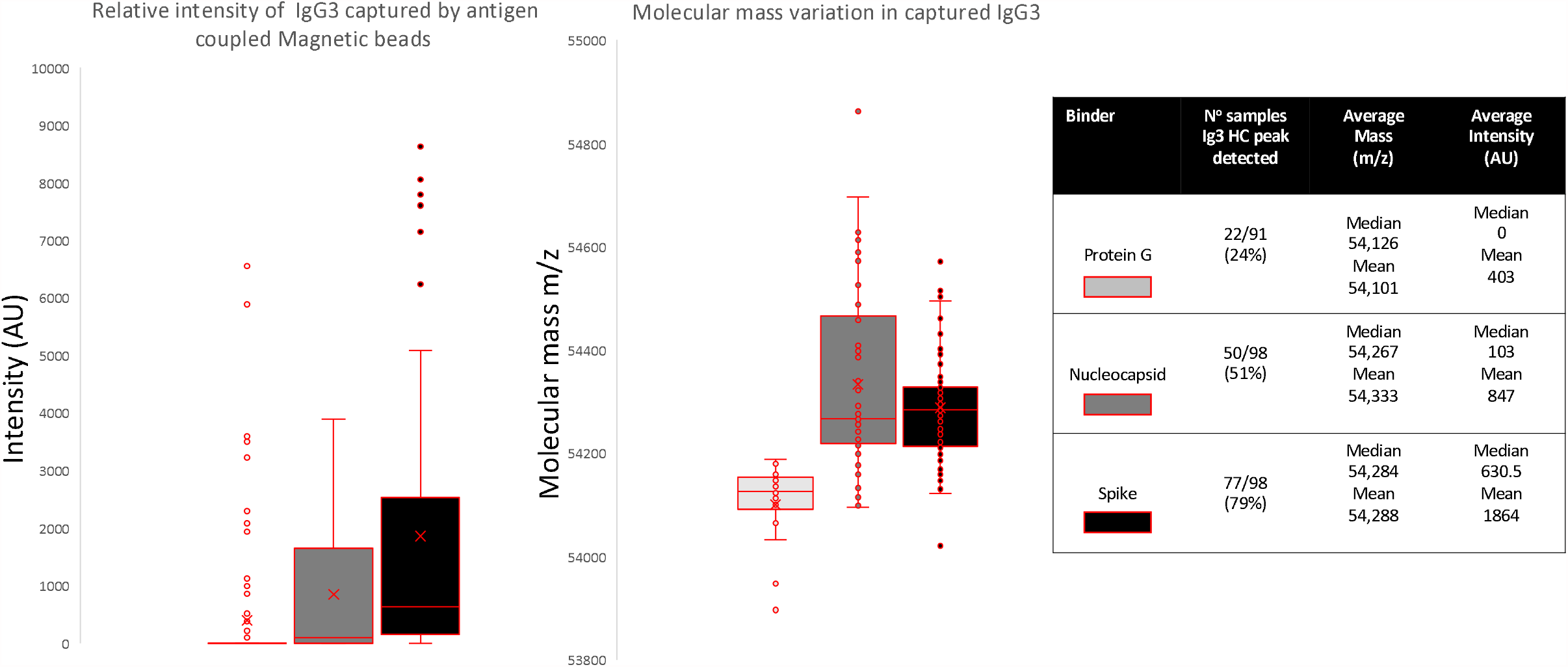
Relative intensities and variance in peak apex molecular mass of IgG3 heavy chains (IgG3 Hc) recovered from the same samples by Protein G, nucleocapsid and stabilised spike protein.

**Figure 5.**
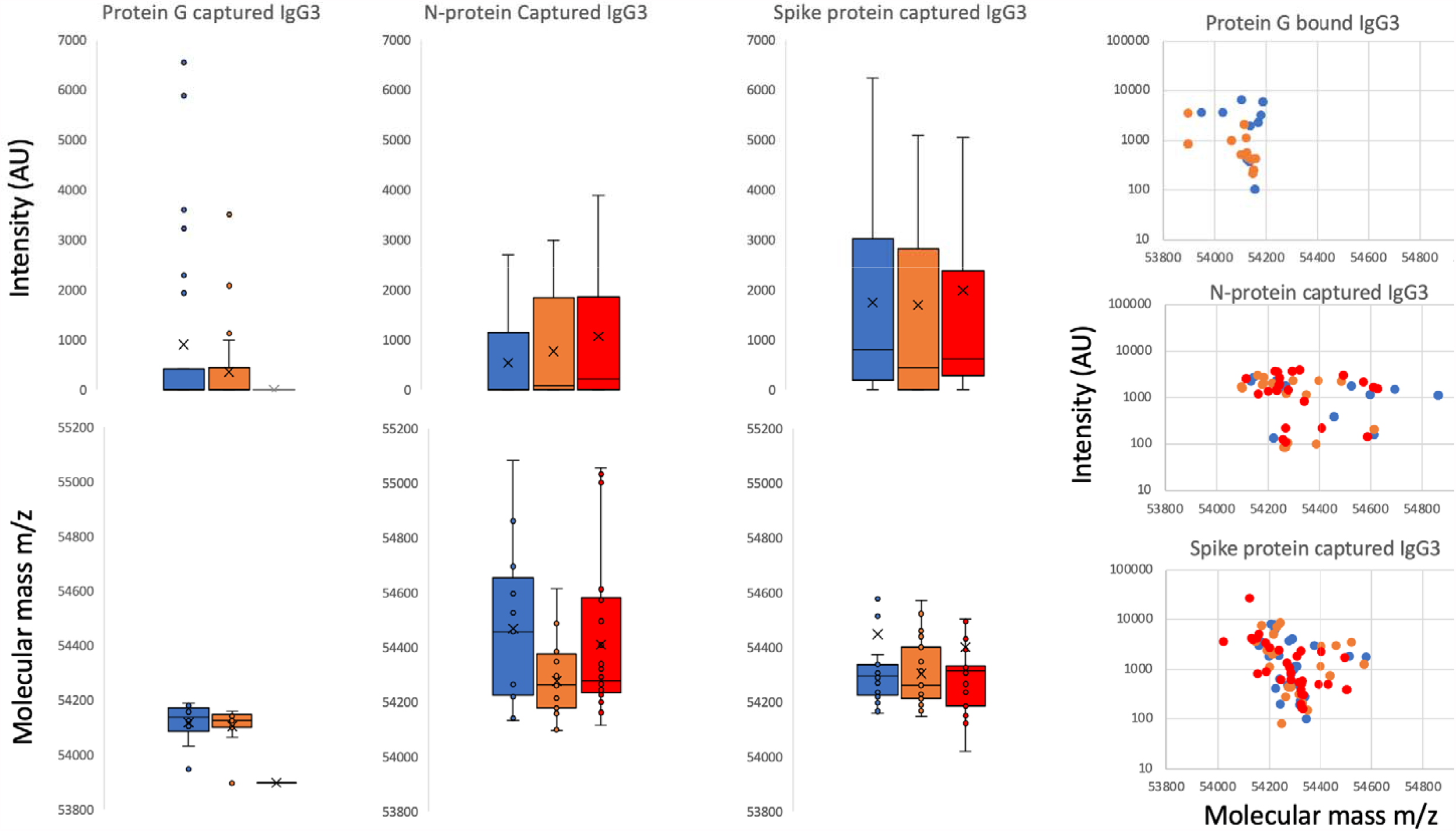
Distribution of intensity for captured and eluted IgG3 heavy chains (IgG3 Hc) and the relative peak molecular mass being bound separated into sample groups. The dot plots to the right are intensity versus molecular mass for individual samples. Blue represents data from SARS-CoV2 sero-negative HCW, Orange from SARS-CoV2 HCW sero-positive having recovered from with mild symptoms and Red sample data from convalescent patients recovering from COVID-19 ARDS.

N-protein showed increased capture of IgG3 by disease severity both in frequency and intensity of binding : sero-negative HCW (peak detected in 51% of samples, median intensity 0AU); sero-positive HCW (peak detected in 52% of samples, median intensity 85AU) and convalescent plasma from COVID-19 ARDS (patients peak detected in 59% of samples, median intensity 221AU). This suggest antibody specific IgG3 binding to N-protein and not a generic binding of total IgG3 by the nucleocapsid protein (see Figures 4&5 and Table 2 panel B). The medium mass of the IgG3 heavy chains were 54,267m/z.

S-protein bound the most IgG3 and this was <u>not</u> associated with history of infection or disease severity: sero-negative HCW (peak detected in 83% of samples, median intensity 889AU); sero-positive HCW (peak detected in 68% of samples, median intensity 443AU) and convalescent plasma from COVID-19 ARDS (patients peak detected in 84% of samples, median intensity 623AU). This reflects immobilised SARS-CoV2 prefusion S-protein having a non-specific binding affinity for IgG3 antibodies, although anti-spike IgG3 specific binding cannot be excluded and would be masked in this system (see Figures 4&5 and Table 2 panel B). A distinctive feature of the IgG3 heavy chains detected in the S-Protein capture experiments was the clear preference for IgG3 with molecular mass of around 54,284 mz (see Figures 4&5 and Table 2 panel B).

## Discussion

The IgG subtypes have unique features associated with complement fixation and Fc receptor binding (see Table1). The two most potent in this respect are IgG1 & IgG3, but IgG3 normally represents only 2-8 % of the Immunoglobulin found in serum and plasma; whilst IgG1 accounts for up to 70%. Here IgG3 was evident as the more dominant IgG subtype in the humoral response to these SARS-COV2 antigens, along with another as yet unidentified Ig with a heavy chain mass of ∼49,000m/z that could be IgG4.

The determination as to which Ig isotype is favoured during maturation of an immune response is influenced in part by cytokine stimulation of the germinal B-cell during Ig heavy chain switching from IgM. Plasma cell formation is induced by IL-21 but modulated by additional cytokines, such as IL-4 promoted switching to IgG; inhibited switching to IgA. Conversely, IL-10 stimulates IgA production [18].

IgG3 has been reported as the dominant antibody in many viral infections [20]. Thus, it is perhaps unsurprising that IgG3 was the dominant captured subtype of IgG in this study. Interestingly, the nucleocapsid protein had a dominant IgG3 humoral patient response whilst the spike also induced a clearly strong IgG1 humoral patient response.

However, unlike the pattern of antibody binding to solid phase antigen seen for IgG1, the prefusion stabilised spike protein also appeared to have a non-specific binding affinity for IgG3, We observed binding of IgG3 from plasma from sero-negative HCW as well as that from patients and sero-positive HCW to S protein. This was similar in this respect to our previously reported affinity of the prefusion S-protein for human serum albumin (HSA) specifically that for a higher mass form due to advanced glycation end product modification; i.e., glycated albumin [16].

The non-specific IgG3 capture by spike protein also appeared to be of a higher mass than the low levels of IgG3 captured from the same samples by Protein-G (Figure 1). Indeed, Protein-G did not bind this higher molecular mass IgG3 found in the COVID-19 ARDS convalescent patients’ samples, despite these being the dominated IgG3 molecular mass form captured by the prefusion S-protein.

The mass differences represent only a 0.4% increase of these Ig Hc, but this would fit with an evasion-pathology hypothesis that SARS COV-2 binds serum proteins with specific glycan residues or reactive glycation end products [16]. Although immunoglobulins can be similarly AGE glycated as a result of elevated blood levels of reducing sugars [20]; there is also the strong possibility that specific variant/ inherent glycosylation of IgG3 is the molecular target of this IgG3 binding-coating by prefusion spike protein. Changes in fucosylation and galactosylation of IgG heavy chain Asn-linked glycan has been reported to be a feature of the humoral immune response of those developing ARDS as a result of COVID-19 [21-23]. However, there is little if any information concerning O-linked glycosylation variation and its effects on antibody bioactivity. IgG3 is unique amongst the IgG subtypes because it has three conserved O-linked glycosylation sites within each of its two defining heavy chain peptides that comprise the extended neck region (see Table 1) thus providing opportunity for great diversity in Fc character [24].

Previous studies in mice and humans suggest that different IgG subclasses show subclass-specific glycosylation patterns [25-27]. In particular human IgG3 had less stem fucosylation and branch terminal galactose Asn-linked glycan moieties than IgG1 [28]. A bias towards IgG3 over IgG1 antibodies against the RBD of spike has been reported as associated with poor prognosis in COVID-19 ARDS patients [12]. Thus, the recent reports of reduced fucose and galactose saccharide residues in anti-SARS-CoV2 IgG N-linked glycans isolated from patients with severe COVID-19 symptoms [22], could be explained by a change in the ratio of IgG3 to IgG1 antibodies. However, the affinity for a specific higher mass form of IgG3 also points to a different post-translational modification, be it hyper glycosylation, glycation or a mixture of the two processes, is associated with ARDS arising in individuals infected with SARS-CoV2.

This may not only prove to be a molecular marker of ARDS susceptibility in COVID-19 infected individuals, but also be directly related to molecular mechanisms by which SARS-CoV-2 can cause the various vascular and immunological pathologies described [29,30].

### Conclusion

The prefusion spike protein of SARS CoV-2 has a binding affinity for serum IgG3 along with HSA which could be mediated via glycation and or variance in inherent glycosylation. This may be part of an immune evasion – misdirection mechanism. The precise nature of the glycation – glycosylation profile in the susceptibility to, and pathogenesis of, COVID-19 ARDS require further study. In addition, humoral immune response reactivity indicate that N-protein induces a more dominant IgG3 response whilst spike protein induces both an IgG1 and IgG3 response. The ratio of IgG1 to IgG3 has been reported by Yates *et al*., [12] to be important in the development of ARDS and this also needs further investigation.

## Data Availability

Upon request to corresponding author

## Abbreviations

HCW: Health care workers
ARDS: Acute respiratory distress syndrome
ITU: Intensive Therapy unit
MALDI-ToF: Matrix assisted laser desorption ionization – time of flight

## Acknowledgements

This study was undertaken by the Humoral Immune Correlates to COVID-19 (HICC) consortium, funded by the UKRI and NIHR; grant number G107217 (COV0170 - HICC: Humoral Immune Correlates for COVID-19). RKI is also funded by NISAD Ideell Förening (charitable association) Organisationsnummer 802528-6157

We gratefully acknowledge Dr Jernej Ule for the loan of a prototype magnetic bead processing rack developed at the Francis Crick Institute London, UK; Bruker UK Ltd, Coventry for the loan of Microflex MALDI-Tof mass spectrometer and Dr Erika Tranfield and Dr Julie Green for technical support with running and data export from the Bruker Microflex.

## References

1. Welker C, Huang J, Gil IJN, Ramakrishna H. Acute Respiratory Distress Syndrome Update, With Coronavirus Disease 2019 Focus J Cardiothorac Vasc Anesth. 2021;S1053-0770(21)00188-9. doi:10.1053/j.jvca.2021.02.053

2. Khadke, S., Ahmed, N., Ahmed, N. et al. Harnessing the immune system to overcome cytokine storm and reduce viral load in COVID-19: a review of the phases of illness and therapeutic agents. Virol J 2020 17, 154 https://doi.org/10.1186/s12985-020-01415-w

3. Cao, X., COVID-19: immunopathology and its implications for therapy. Nature reviews immunology, 2020 20(5), 269–270.

4. Wang W, Xu Y, Gao R, et al. Detection of SARS-CoV-2 in Different Types of Clinical Specimens. JAMA.2020;323(18):1843–1844. doi:10.1001/jama.2020.3786

5. Azkur, A.K., Akdis, M., Azkur, D., S et al. Immune response to SARSDCoVD2 and mechanisms of immunopathological changes in COVIDD19. Allergy, 2020 75(7), pp.1564–1581.

6. Post, N., Eddy, D., Huntley, C. et al.. Antibody response to SARS-CoV-2 infection in humans: A systematic review. PloS one, 2020 15(12), p.e0244126.

7. Galipeau, Y., Greig, M., Liu, G., Driedger, M. and Langlois, M.A., 2020. Humoral Responses and Serological Assays in SARS-CoV-2 Infections. Frontiers in immunology, 2020 11.

8. Voss C, Esmail S, Liu X, Knauer MJ, et al. Epitope-specific antibody responses differentiate COVID-19 outcome and variants of concern. JCI Insight. 2021 :148855. doi: 10.1172/jci.insight.148855.

9. Robbiani DF, Gaebler C, Muecksch F, et al. Convergent antibody responses to SARS-CoV-2 in convalescent individuals. Nature. 2020 584(7821):437–442. doi: 10.1038/s41586-020-2456-9.

10. Suthar MS, Zimmerman MG, Kauffman et al. Rapid Generation of Neutralizing Antibody Responses in COVID-19 Patients. Cell Rep Med. 1(3):100040. doi: 10.1016/j.xcrm.2020.100040.

11. Atyeo C, Fischinger S, Zohar T, et al.. Distinct Early Serological Signatures Track with SARS-CoV-2 Survival. Immunity 2020 15;53(3):524-532.e4. doi: 10.1016/j.immuni.2020.07.020.

12. Jennifer L. Yates, Dylan J. Ehrbar, Danielle T. Hunt, et al. Serological Analysis Reveals an Imbalanced IgG Subclass Composition Associated with COVID-19 Disease Severity medRxiv 2020.10.07.20208603; doi:https://doi.org/10.1101/2020.10.07.20208603

13. Castillo-Olivares, J., Wells, D.A., Ferrari, et al. Towards Internationally standardised humoral Immune Correlates of Protection from SARS-CoV-2 infection and COVID-19 disease. medRxiv 2021.05.21.21257572;:https://doi.org/10.1101/2021.05.21.21257572

14. Song W, Gui M, Wang X, Xiang Y (2018) Cryo-EM structure of the SARS coronavirus spike glycoprotein in complex with its host cell receptor ACE2. PLoS Pathog 14(8): e1007236. https://doi.org/10.1371/journal.ppat.1007236

15. Carnell, G.W., Ciazynska, K.A., Wells, D.A., Xiong, X., Aguinam, E.T., McLaughlin, S.H., Mallery, D., Ebrahimi, S., Ceron-Gutierrez, L., Asbach, B. and Einhauser, S., 2021. SARS-CoV-2 spike protein stabilized in the closed state induces potent neutralizing responses. Journal of virology, pp.JVI-00203.

16. Iles JK, Zmuidinaite, R Saddee, C et al. SARS-CoV-2 Spike protein binding of glycated serum albumin -its potential role in the pathogenesis of the COVID-19 clinical syndromes and bias towards individuals with pre-diabetes/type 2 diabetes & metabolic diseases. medRxiv 2021.06.14.21258871; doi:https://doi.org/10.1101/2021.06.14.21258871

17. Strohalm, M., Kavan, D., Novak, P., Volny, M. and Havlicek, V., mMass 3: a cross-platform software environment for precise analysis of mass spectrometric data. Analytical chemistry, 2010 82, 4648–4651.

18. Lucas, C.L., Kuehn, H.S., Zhao, F., et al. Dominant-activating germline mutations in the gene encoding the PI (3) K catalytic subunit p110 δ result in T cell senescence and human immunodeficiency. Nature immunology, 2014 15(1), 88–97.

19. Walker, M.R., Eltahla, A.A., Mina, M.M., Li, H., Lloyd, A.R., Bull, R.A. Envelope-Specific IgG3 and IgG1 Responses Are Associated with Clearance of Acute Hepatitis C Virus Infection. Viruses 2020, 12, 75. https://doi.org/10.3390/v12010075

20. Pampati PK, Suravajjala S, Dain JA. Monitoring nonenzymatic glycation of human immunoglobulin G by methylglyoxal and glyoxal: A spectroscopic study. Anal Biochem. 2011;408(1):59–63. doi:10.1016/j.ab.2010.08.038

21. Hoepel, W., Chen, H.J., Allahverdiyeva, S. et al. 2020. High titers and low fucosylation of human anti–spike protein IgG promote alveolar macrophage activation and inflammation. Science Translational Medicine 2021:13,596,eabf8654 DOI: 10.1126/scitranslmed.abf8654

22. Larsen, M.D., de Graaf, E.L., Sonneveld, M.E. et al. Afucosylated IgG characterizes enveloped viral responses and correlates with COVID-19 severity. Science 2021, 371(6532).

23. Chakraborty, S., Gonzalez, J., Edwards, K., et al. Proinflammatory IgG Fc structures in patients with severe COVID-19. Nature Immunology, 2021 22(1), 67–73.

24. Plomp R, Dekkers G, Rombouts Y, et al. Hinge-Region O-Glycosylation of Human Immunoglobulin G3 (IgG3). Mol Cell Proteomics. 2015 14(5):1373–1384. doi:10.1074/mcp.M114.047381

25. Kao, D., Lux, A., Schaffert, A., Lang, R., Altmann, F., and Nimmerjahn, F. IgG subclass and vaccination stimulus determine changes in antigen specific antibody glycosylation in mice. Eur. J. Immunol. 2017 47, 2070–2079. doi: 10.1002/eji.201747208

26. de Haan, N., Reiding, K. R., Krišti ć, J., Hipgrave Ederveen, A. L., Gordan, L., and Manfred, W. The N-glycosylation of mouse immunoglobulin g (IgG) -Fragment crystallizable differs between igg subclasses and strains. Front. Immunol. 2017 8:608. doi: 10.3389/fimmu.2017.00608

27. Zaytseva, O. O., Jansen, B. C., Haniæ, M., et al. MIgGGly (mouse IgG glycosylation analysis) - a high-throughput method for studying Fc-linked IgG N-glycosylation in mice with nanoUPLC-ESI-MS. Sci. Rep. 2018 8:13688. doi: 10.1038/s41598-018-31844-31841

28. Wieczorek M, Braicu EI, Oliveira-Ferrer Le, Sehouli J, and Blanchard V. Immunoglobulin G Subclass-Specific Glycosylation Changes in Primary Epithelial Ovarian Cancer Frontiers in Immunology 2020 11:654 DOI=10.3389/fimmu.2020.00654

29. Costa LB, Perez LG, Palmeira VA, et al. AC. Insights on SARS-CoV-2 Molecular Interactions With the Renin-Angiotensin System. Front Cell Dev Biol. 2020 8:559841. doi: 10.3389/fcell.2020.559841.

30. Sood, S., Aggarwal, V., Aggarwal, D. et al. COVID-19 Pandemic: from Molecular Biology, Pathogenesis, Detection, and Treatment to Global Societal Impact. Curr Pharmacol Rep 2020 6, 212–227. https://doi.org/10.1007/s40495-020-00229-2

